# Importance of Interaction Structure and Stochasticity for Epidemic Spreading: A COVID-19 Case Study

**DOI:** 10.1101/2020.05.05.20091736

**Authors:** Gerrit Großmann, Michael Backenköhler, Verena Wolf

## Abstract

In the recent COVID-19 pandemic, computer simulations are used to predict the evolution of the virus propagation and to evaluate the prospective effectiveness of non-pharmaceutical interventions. As such, the corresponding mathematical models and their simulations are central tools to guide political decision-making. Typically, ODE-based models are considered, in which fractions of infected and healthy individuals change deterministically and continuously over time.

In this work, we translate an ODE-based COVID-19 spreading model from literature to a stochastic multi-agent system and use a contact network to mimic complex interaction structures. We observe a large dependency of the epidemic’s dynamics on the structure of the underlying contact graph, which is not adequately captured by existing ODE-models. For instance, existence of super-spreaders leads to a higher infection peak but a lower death toll compared to interaction structures without super-spreaders. Overall, we observe that the interaction structure has a crucial impact on the spreading dynamics, which exceeds the effects of other parameters such as the basic reproduction number *R*_0_.

We conclude that deterministic models fitted to COVID-19 outbreak data have limited predictive power or may even lead to wrong conclusions while stochastic models taking interaction structure into account offer different and probably more realistic epidemiological insights.

## 1 Introduction

On March 11th, 2020, the World Health Organization (WHO) officially declared the outbreak of the *coronavirus disease* 2019 (COVID-19) to be a pandemic. By this date at the latest, curbing the spread of the virus became a major worldwide concern. Given the lack of a vaccine, the international community relied on non-pharmaceutical interventions (NPIs) such as social distancing, mandatory quarantines, or border closures. Such intervention strategies, however, inflict high costs on society. Hence, for political decision-making it is crucial to forecast the spreading dynamics and to estimate the effectiveness of different interventions.

Mathematical and computational modeling of epidemics is a long-established research field with the goal of predicting and controlling epidemics. It has developed epidemic spreading models of many different types: data-driven and mechanistic as well as deterministic and stochastic approaches, ranging over many different temporal and spatial scales (see [50,15] for an overview).

Computational models have been calibrated to predict the spreading dynamics of the COVID-19 pandemic and influenced public discourse. Most models and in particular those with high impact are based on ordinary differential equations (ODEs). In these equations, the fractions of individuals in certain compartments (e.g., infected and healthy) change continuously and deterministically over time, and interventions can be modeled by adjusting parameters.

In this paper, we compare the results of COVID-19 spreading models that are based on ODEs to results obtained from a different class of models: stochastic spreading processes on contact networks. We argue that virus spreading models taking into account the interaction structure of individuals and reflecting the stochasticity of the spreading process yield a more realistic view on the epidemic’s dynamics.

If an underlying interaction structure is considered, not all individuals of a population meet equally likely as assumed for ODE-based models. A well-established way to model such structures is to simulate the spreading on a network structure that represents the individuals of a population and their social contacts. Effects of the network structure are largely related to the *epidemic threshold* which describes the minimal *infection rate* needed for a pathogen to be able to spread over a network [38]. In the network-free paradigm the basic reproduction number (*R*_0_), which describes the (mean) number of susceptible individuals infected by patient zero, determines the evolution of the spreading process. The value *R*_0_ depends on both, the connectivity of the society *and* the infectiousness of the pathogen. In contrast, in the network-based paradigm the interaction structure (given by the network) and the infectiousness (given by the infection rate) are decoupled.

Here, we focus on contact networks as they provide a universal way of encoding real-world interaction characteristics like super-spreaders, grouping of different parts of the population (e.g. senior citizens or children with different contact patterns), as well as restrictions due to spatial conditions and mobility, and household structures. Moreover, models based on contact networks can be used to predict the efficiency of interventions [39,35,5].

Here, we analyze in detail a network-based stochastic model for the spreading of COVID-19 with respect to its differences from existing ODE-based models and the sensitivity of the spreading dynamics on particular network features. We calibrate both, ODE-models and stochastic models with interaction structure to the same basic reproduction number *R*_0_ or to the same infection peak and compare the corresponding results. In particular, we analyze the changes in the effective reproduction number over time. For instance, early exposure of superspreaders leads to a sharp increase of the reproduction number, which results in a strong increase of infected individuals. We compare the times at which the number of infected individuals is maximal for different network structures as well as the death toll. Our results show that the interaction structure has a major impact on the spreading dynamics and, in particular, important characteristic values deviate strongly from those of the ODE model.

## 2 Related Work

In the last decade, research focused largely on epidemic spreading, where interactions were constrained by contact networks, i.e., a graph representing the individuals (as nodes) and their connectivity (as edges). Many generalizations, e.g. to weighted, adaptive, temporal, and multi-layer networks exist [32,45]. Here, we focus on simple contact networks without such extensions.

Spreading characteristics on different contact networks based on the Susceptible-Infected-Susceptible (SIS) or Susceptible-Infected-Recovered (SIR) compartment model have been investigated intensively. In such models, each individual (node) successively passes through the individual stages (compartments). For an overview, we refer the reader to [36]. Qualitative and quantitative differences between network structures and network-free models have been investigated in [23,2]. In contrast, this work considers a specific COVID-19 spreading model and focuses on those characteristics that are most relevant for COVID-19 and which have, to the best of our knowledge, not been analyzed in previous work.

SIS-type models require knowledge of the spreading parameters (infection strength, recovery rate, etc.) and the contact network, which can partially be inferred from real-world observations. Currently, inferred data for COVID-19 seems to be of very poor quality [25]. However, while the spreading parameters are subject to a broad scientific discussion. Publicly available data, which could be used for inferring a realistic contact network, practically does not exist. Therefore real-world data on contact networks is rare [31,46,24,33,44] and not available for large-scale populations. A reasonable approach is to generate the data synthetically, for instance by using mobility and population data based on geographical diffusion [47,18,37,3]. For instance, this has been applied to the influenza virus [34]. Due to the major challenge of inferring a realistic contact network, most of these works, however, focus on how specific network features shape the spreading dynamics.

### 2.1 COVID-19 Spreading Models

Literature abounds with proposed models of the COVID-19 spreading dynamics. Very influential is the work of Neil Ferguson and his research group that regularly publishes reports on the outbreak (e.g. [11]). They study the effects of different interventions on the outbreak dynamics. The computational modeling is based on a model of influenza outbreaks [20,12]. They present a very high-resolution spatial analysis based on movement-data, air-traffic networks etc. and perform sensitivity analysis on the spreading parameters, but to the best of our knowledge not on the interaction data. Interaction data were also inferred locally at the beginning of the outbreak in Wuhan [4] or in Singapore [41] and Chicago [13]. Models based on community structures, however, consider isolated (parts of) cities and are of limited significance for large-scale model-based analysis of the outbreak dynamic.

Another work focusing on interaction structure is the modeling of outbreak dynamics in Germany and Poland done by Bock et al. [6]. The interaction structure within households is modeled based on census data. Inter-household interactions are expressed as a single variable and are inferred from data. They then generated “representative households” by re-sampling but remain vague on many details of the method.

A more rigorous model of stochastic propagation of the virus is proposed by Arenas et al. [1]. They take the interaction structure and heterogeneity of the population into account by using demographic and mobility data. They analyze the model by deriving a mean-field equation. Mean-field equations are more suitable to express the mean of a stochastic process than other ODE-based methods but tend to be inaccurate for complex interaction structures. Moreover, the relationship between networked-constrained interactions and mobility data remains unclear to us.

Other notable approaches use SIR-type methods, but cluster individuals into age-groups [40,29], which increases the model’s accuracy. Rader et al. [42] combined spatial-, urbanization-, and census-data and observed that the crowding structure of densely populated cities strongly shaped the epidemics intensity and duration. In a similar way, a meta-population model for a more realistic interaction structure has been developed [8] without considering an explicit network structure.

The majority of research, however, is based on deterministic, network-free SIR-based ODE-models. For instance, the work of Jose Lourenco et al. [30] infers epidemiological parameters based on a standard SIR model. Similarly, Dehning et al. [9] use an SIR-based ODE-model, where the infection rate may change over time. They use their model to predict a suitable time point to loosen NPIs in Germany. Khailaie et al. analyze how changes in the reproduction number (“mimicking NPIs”) affect the epidemic dynamics [26], where a variant of the deterministic, network-free SIR-model is used and modified to include states (compartments) for hospitalized, deceased, and asymptomatic patients. Otherwise, the method is conceptually very similar to [30,9] and the authors argue against a relaxation of NPIs in Germany. Another popular work is the online simulator *covidsim*^1^. The underlying method is also based on a network-free SIR-approach [51,52]. However, the role of an interaction structure is not discussed and the authors explicitly state that they believe that the stochastic effects are only relevant in the early stages of the outbreak. A very similar method has been developed at the German Robert-Koch-Institut (RKI) [7]. Jianxi Luo et al. proposed an ODE-based SIR-model to predict the end of the COVID-19 pandemic^2^, which is regressed with daily updated data. ODE-models have also been used to project the epidemic dynamics into the “postpandemic” future by Kissler et al. [28]. Some groups also resort to to branching processes, which are inherently stochastic but not based on a complex interaction structure [22,43].

**Fig. 1:**
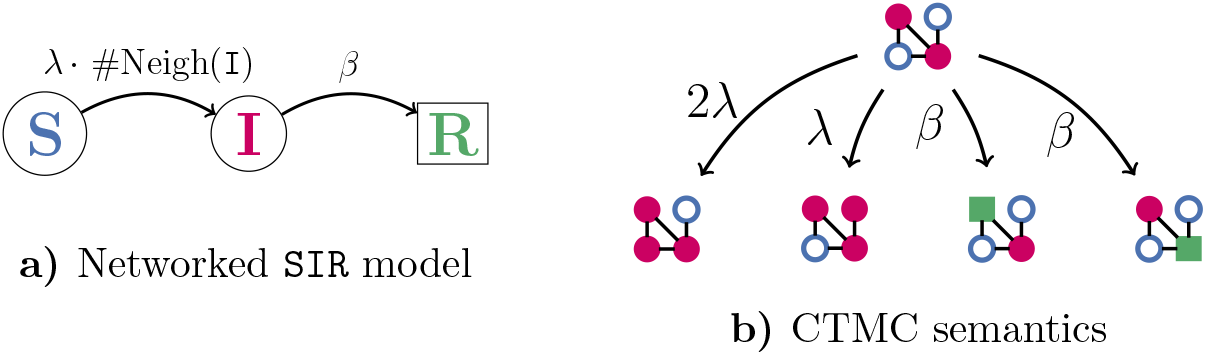
Networked SIR model. (a) Compartments with instantaneous transition rates. Each node successively passes through the three compartments/states: susceptible (S), infected (I), and recovered/removed (R). (b) Four possible transitions on a 4-node contact network based on the CTMC semantics.

## 3 Translating SIR-type Models for Epidemic Spreading

A very popular class of epidemic models is based on the assumption that during an epidemic individuals are either susceptible (S), infected (I), or recovered/removed (R). The mean number of individuals in each compartment evolves according to the following system of ordinary differential equations

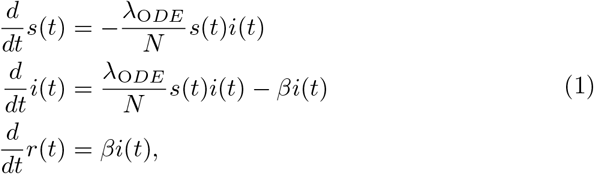

where *N* denotes the total population size, λ*_ODE_* and *β* are the infection and recovery rates. Typically, one assumes that *N* = 1 in which case the equation refers to fractions of the population, leading to the invariance *s*(*t*)*+i*(*t*)*+r*(*t*) = 1 for all t. It is trivial to extend the compartments and transitions.

### 3.1 Network-Based Spreading Model

A stochastic network-based spreading model is a continuous-time stochastic process on a discrete state space. The underlying structure is given by a graph, where each node represents one individual (or any other entity of interest). At each point in time, each node occupies a compartment, for instance: S, I, or R. Moreover, nodes can only receive or transmit infections from neighboring nodes (according to the edges of the graph). For the general case with m possible compartments, this yields a state space of size *m^n^*, where n is the number of nodes. The jump times until events happen are typically assumed to follow an exponential distribution. Note that in the ODE model, residual residence times in the compartments are not tracked, which naturally corresponds to the exponential distribution in the network model. Hence, the underlying stochastic process is a continuous-time Markov Chain (CTMC) [27]. The extension to non-Markovian semantics is trivial. We illustrate the three-compartment case in Fig. 1. The transition rates of the CTMC are such that an infected node transmits infections at rate λ. Hence, the rate at which a susceptible node is infected is λ-#Neigh(I), where #Neigh(I) is the number of its infected direct neighbors. Spontaneous recovery of a node occurs at rate *β*. The size of the state space renders a full solution of the model infeasible and approximations of the mean-field [14] or Monte-Carlo simulations are common ways to analyze the process.

#### General Differences to the ODE model

The aforementioned formalism yields some fundamental differences from network-free ODE-based approaches. The most distinct difference is the decoupling of infectiousness and interaction structure. The infectiousness λ (i.e., the infection rate) is assumed to be a parameter expressing how contagious a pathogen inherently is. It encodes the probability of a virus transmission *if* two people meet. That is, it is independent from the social interactions of individuals (it might however depend on hygiene, masks, etc.). The influence of social contacts is expressed in the (potentially time-varying) connectivity of the graph. Loosely speaking, it encodes the possibility *that* two individuals meet. In the ODE-approach both are combined in the basic reproduction number. Note that, throughout this manuscript, we use λ to denote the infectiousness of COVID-19 (as an instantaneous transmission rate).

Another important difference is that ODE-models consider fractions of individuals in each compartment. In the network-based paradigm, we model absolute numbers of entities in each compartment and extinction of the epidemic may happen with positive probability. While ODE-models are agnostic to the actual population size, in network-based models, increasing the population by adding more nodes inevitably changes the dynamics.

A key link between the two paradigms is that if the network topology is a *complete graph* (resp. clique) then the ODE-model gives an accurate approximation of the expected fractions of the network-based model. In systems biology this assumption is often referred to as *well-stirredness*. In the limit of an infinite graph size, the approximation approaches the true mean.

### 3.2 From ODE-Models to Networks

To transform an ODE-model to a network-based model, one can simply keep rates relating to spontaneous transitions between compartments as these transitions do not depend on interactions (e.g., recovery at rate *β*). Translating the infection rate is more complicated. In ODE-models, one typically has given an infection rate and assumes that each infected individual can infect all susceptible ones. To make the model invariant to the actual number of individuals, one typically divides the rate by the population size (or assumes the population size is one and the ODEs express fractions). Naturally, in a contact network, we do not work with fractions but each node relates to one entity.

Here, we propose to choose an infection rate such that the network-based model yields the same basic reproduction number *R*_0_ as the ODE-model. The basic reproduction number describes the (expected) number of individuals that an infected person infects in a completely susceptible population. We calibrate our model to this starting point of the spreading process, where there is a single infected node (*patient zero*). We assume that *R*_0_ is either explicitly given or can implicitly be derived from an ODE-based model specification. Hence, when we pick a random node as patient zero, we want it to infect on average *R*_0_ susceptible neighbors (all neighbors are susceptible at that point in time) before it recovers or dies.

Let us assume that, like in the aforementioned SIR-model, infectious node infect their susceptible neighbors with rate λ and that an infectious node loses its infectiousness (by dying, recovering, or quarantining) with rate *β*. According to the underlying CTMC semantics of the network model, each susceptible neighbor gets infected with probability 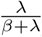 [27]. Note that we only take direct infections from patient zero into account and, for simplicity, assume all neighbors are only infected by patient zero. Hence, when patient zero has *k* neighbors, the expected number of neighbors it infects is 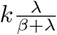. Since the mean degree of the network is *k*_mean_, the expected number of nodes infected by patient zero is

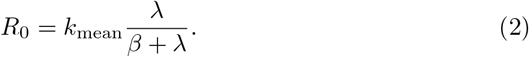

Now we can calibrate λ to relate to any desired *R*_0_. That is

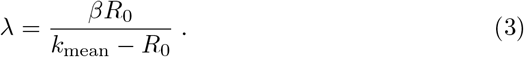

Note that *R*0 will always be smaller than *k*_mean_ which follows from Eq. (2), considering that *k*_mean_ ≥1 (by construction of the network) and *β* > 0. In contrast, in the deterministic paradigm this relationship is given by the equation (cf. [30,9]):

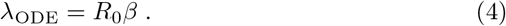

Note that the recovery rate *β* is identical in the ODE-and network-model. We can translate the infection rate of an ODE-model to a corresponding network-based stochastic model with the equation

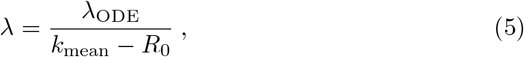

while keeping *R*_0_ fixed. In the limit of an infinite complete network, this yields 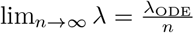, which is equivalent to the effective infection rate in the ODE-model 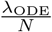 for population size *N* (cf. Eq. (1)).

#### Example

Consider a network where each node has exactly 5 neighbors (a 5-regular graph) and let *R*_0_ = 2. We also assume that the recovery rate is *β* = 1, which then yields Aode = 2. The probability that a random neighbor of patient zero becomes infected is 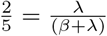, which gives 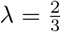.

#### Extensions of SIR

It is trivial to extent the compartments and transitions, for instance by including an *exposed* compartment for the time-period where an individual is infected but not yet infectious. The derivation of R_0_ remains the same. The only requirement is the existence of a distinct infection and recovery rate, respectively. In the next section, we discuss a more complex case.

## 4 A Network-based COVID-19 Spreading Model

We consider a network-based model that is strongly inspired by the ODE-model used in [26] and document it in Fig. 2. We use the same compartments and transition-types but simplify the notation compared to [26] to make the intuitive meaning of the variables clearer.^3^

We denote the compartments by 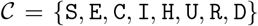, where each node can be *susceptible* (S), *exposed* (E), a *carrier* (C), *infected* (I), *hospitalized* (H), in the *intensive care unit* (U), *dead* (D), or *recovered* (R). Exposed agents are already infected but symptom-free and not infectious. Carriers are also symptom-free but already infectious. Infected nodes show symptoms and are infectious. Therefore, we assume that their infectiousness is reduced by a factor of *γ* (*γ* ≤ 1, sick people will reduce their social activity). Individuals that are hospitalized (or in the ICU) are assumed to be properly quarantined and cannot infect others.

Accurate spreading parameters are very difficult to infer in general and the high number of undetected cases complicates the problem further in the current pandemic. Here, we choose values that are within the ranges listed in [26], where the ranges are rigorously discussed and justified. We document them in Table 1. We remark that there is a high amount of uncertainty in the spreading parameters. However, our goal is not a rigorous fit to data but rather a comparison of network-free ODE-models to stochastic models with an underlying network structure.

Note that the mean number of days in a compartment is the inverse of the cumulative instantaneous rate to leave that compartment. For instance, the mean residence time in compartment H is 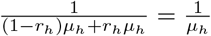. As a consequence of the race condition of the exponential distribution [48], *r_h_* modulates the probability of entering the successor compartment. That is, with probability *r_h_*, the successor compartment will be R and not U.

Inferring the infection rate λ for a fixed *R*_0_ is somewhat more complex than in the previous section because this model admits two compartments for infectious agents. We first consider the expected number of nodes that a randomly chosen patient zero infects, while being in state C. We denote the corresponding basic reproduction number by 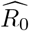. We calibrate the only unknown parameter λ accordingly (the relationships from the previous section remain valid). We explain the relation to *R*_0_ when taking C and I into account in the Appendix (available at [16]). Substituting *β* by *μ_c_* gives

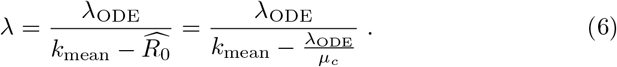

### 4.1 Human-to-Human Contact Networks

Naturally, it is extremely challenging to reconstruct large-scale contact-networks based on data. Here, we test different types of contact networks with different features, which are likely to resemble important real-world characteristics. The contact networks are specific realizations (i.e., variates) of random graph models. Different graph models highlight different (potential) features of the real-world interaction structure. The number of nodes ranges from 100 to 10^5^. We only use strongly connected networks (where each node is reachable from all other nodes). We refer to [10] or the NetworkX [19] documentation for further information about the network models discussed in the sequel. We provide a schematic visualization in Fig. 3.

We consider **Erdős–Rényi** (ER) random graphs as a baseline, where each pair of nodes is connected with a certain (fixed) probability. We also compute results for **Watts–Strogatz** (WS) random networks. They are based on a ring topology with random re-wiring. The re-wiring yields to a small-world property of the network. Colloquially, this means that one can reach each node from each other node with a small number of steps (even when the number of nodes increases). We further consider **Geometric Random Networks** (GN), where nodes are randomly sampled in an Euclidean space and randomly connected such that nodes closer to each other have a higher connection probability. We also consider **Barabási–Albert** (BA) random graphs, that are generated using a preferential attachment mechanism among nodes, as well as graphs generated using the **Configuration Model** (CM-PL) which are—except from being constrained on having power-law degree distribution—completely random. The latter two models models contain a very small number of nodes with very high degree, which act as super-spreaders. We also test a synthetically generated **Household** (HH) network that was loosely inspired by [2]. Each household is a clique, the edges between households represent connections stemming from work, education, shopping, leisure, etc. We use a configuration model to generate the global inter-household structure that follows a power-law distribution. We also use a **complete graph** (CG) as a sanity check. It allows the extinction of the epidemic, but otherwise similar results to those of the ODE are expected.

**Fig. 2:**
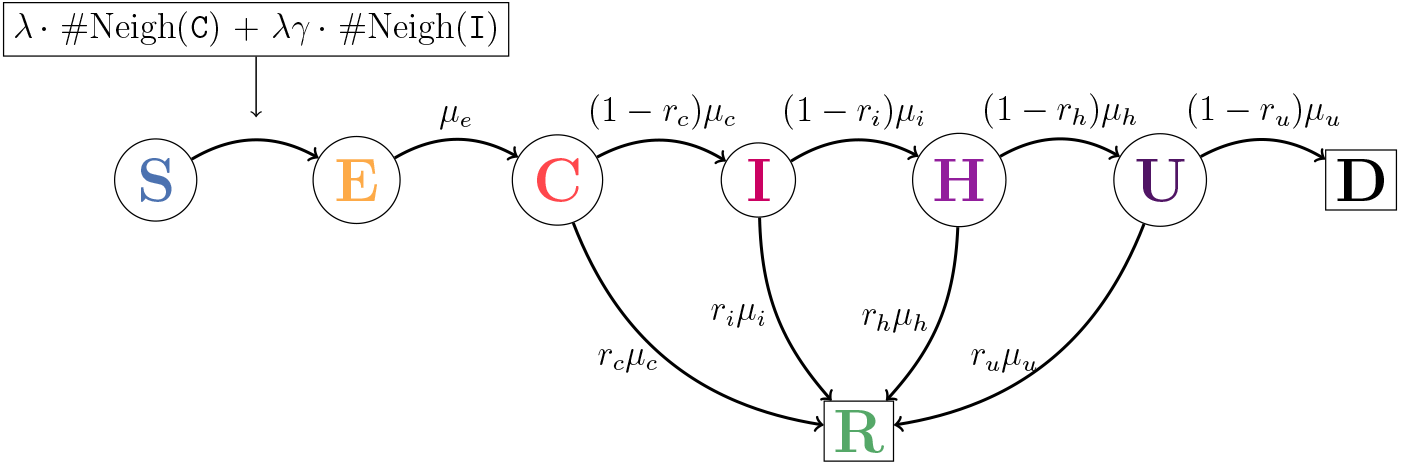
Multi-agent compartment model for COVID-19 with instantaneous transition rates. The infection rate is λ. Exposed (E) nodes are newly infected. Carriers (C) are already infectious but still symptomless, infected nodes (I) develop symptoms and reduce their social activity (modulated by γ). Nodes that are hospitalized (H) and in the ICU (U) are properly quarantined. Recovered (R) nodes remain recovered.

**Table 1:**
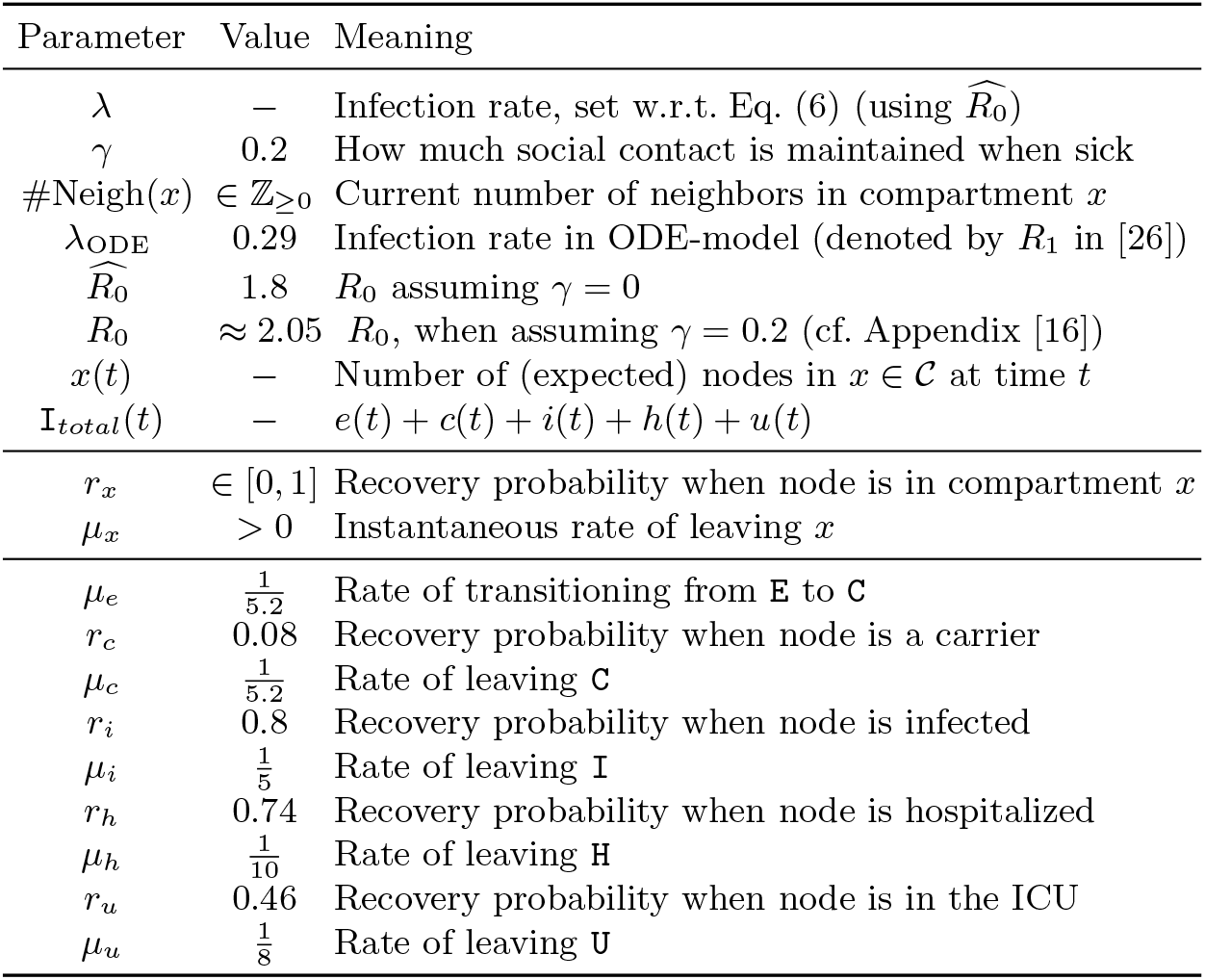
Model Parameters

**Fig. 3:**
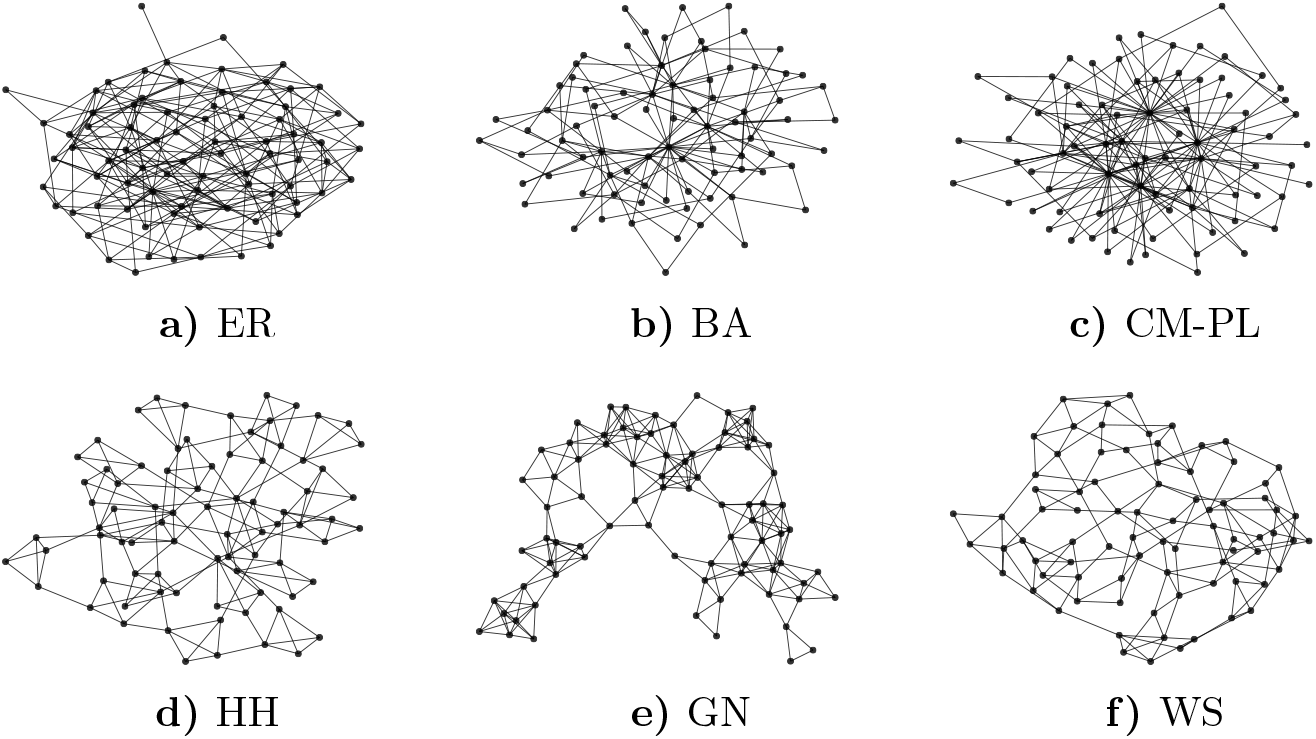
Schematic visualizations of the random graph models with 80 nodes.

### 4.2 Parameter Calibration

We are interested in the relationship between the contact network structure, *R*_0_, the height and time point of the infection-peak, and the number of individuals ultimately affected by the epidemic. Therefore, we run different network models with different 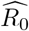. For one series of experiments, we fix 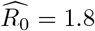 and derive the corresponding infection rate λ and the value for λ_ODE_ in the ODE model. In the second experiments, calibrate λ and λ_ODE_ such that all infection peaks lie on the same level.

### 4.3 Interventions

In the sequel, we do not explicitly model NPIs. However, we note that the network-based paradigm makes it intuitive to distinguish between NPIs related to the probability that people meet (by changing the contact network) and NPIs related to the probability of a transmission happening when two people meet (by changing the infection rate λ). Political decision-making is faced with the challenge of transforming a network structure which inherently supports COVID-19 spreading to one which tends to suppress it. Here, we investigate how changes in λ affect the dynamics of the epidemic in Section 5 (Experiment 3).

## 5 Numerical Results

We compare the solution of the ODE model (using numerical integration) with the solution of the corresponding stochastic network-based model (using MonteCarlo simulations). Code will be made available^4^. We investigate the evolution of mean fractions in each compartment over time, the evolution of the so-called *effective reproduction number*, and the influence of the infectiousness λ.

**Setup**. We used contact networks with n = 1000 nodes (except for the complete graph where we used 100 nodes). To generate samples of the stochastic spreading process, we used event-driven simulation (similar to the rejection-free version in [17]). Specifically, we utilized a simulation scheme, where all future events (i.e., transitions of nodes) were sorted in a priority queue (according to their application time). In each simulation step, the next event is drawn from the queue, the event is applied to the network, and the time is updated accordingly. Depending on the type of transition (a) new event(s) is (are) generated and pushed to the queue. Some events already in the queue might become irrelevant and are removed. The event queue is initialized by generating one event for each node.

The simulation started with three random seed nodes in compartment C (and with an initial fraction of 3/1000 for the ODE model). One thousand simulation runs were performed on a fixed variate of a random graph. We remark that results for other variates were very similar. Hence, for better comparability, we refrained from taking an average over the random graphs. The parameters to generate a graph are: ER: *k*_mean_ = 6, WS: *k* = 4 (numbers of neighbors), *p* = 0.2 (re-wire probability), GN: *r* = 0.1 (radius), BA: m = 2 (number of nodes for attachment), CM-PL: *γ* = 2.0 (power-law parameter), *k*_min_ = 2, HH: household size is 4, global network is CM-PL with *γ* = 2.0, *k*_min_ = 3. The CPU time for a single simulation on a standard desktop computer was in the range of a few hours.

**Experiment 1: Results with Homogeneous 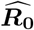**. In our first experiment, we compare the epidemic’s evolution (cf. Fig. 4) while λ is calibrated such that all networks admit an 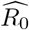 of 1.8. and λ is set (w.r.t. the mean degree) according to Eq. (6). Thereby, we analyze how well certain network structures generally support the spread of COVID-19. The evolution of the mean fraction of nodes in each compartment is illustrated in Fig. 4 and Fig. 5.

Based on the Monte-Carlo simulations, we analyzed how *R_t_*, the *effective reproduction number*, changes over time. The number *R_t_* denotes the average number of neighbors that an infectious node (who got exposed at day t) infects over time (cf. Fig. 6). For *t* = 0, the estimated effective reproduction number always starts around the same value and matched the theoretical prediction. Independent of the network, 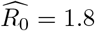 yields *R*_0_ ≈ 2.05 (cf. Appendix [16]).

In Fig. 6 we see that the evolution of *R_t_* differs tremendously for different contact networks. Unsurprisingly, *R_t_* decreases on the complete graph (CG), as nodes, that become infectious later, will not infect more of their neighbors. This also happens for GN-and WS-networks, but they cause a much slower decline of *R_t_* which is around 1 in most parts (the sharp decrease in the end stems from the end of the simulation being reached). This indicates that the epidemic slowly “burns” through the network.

**Fig. 4:**
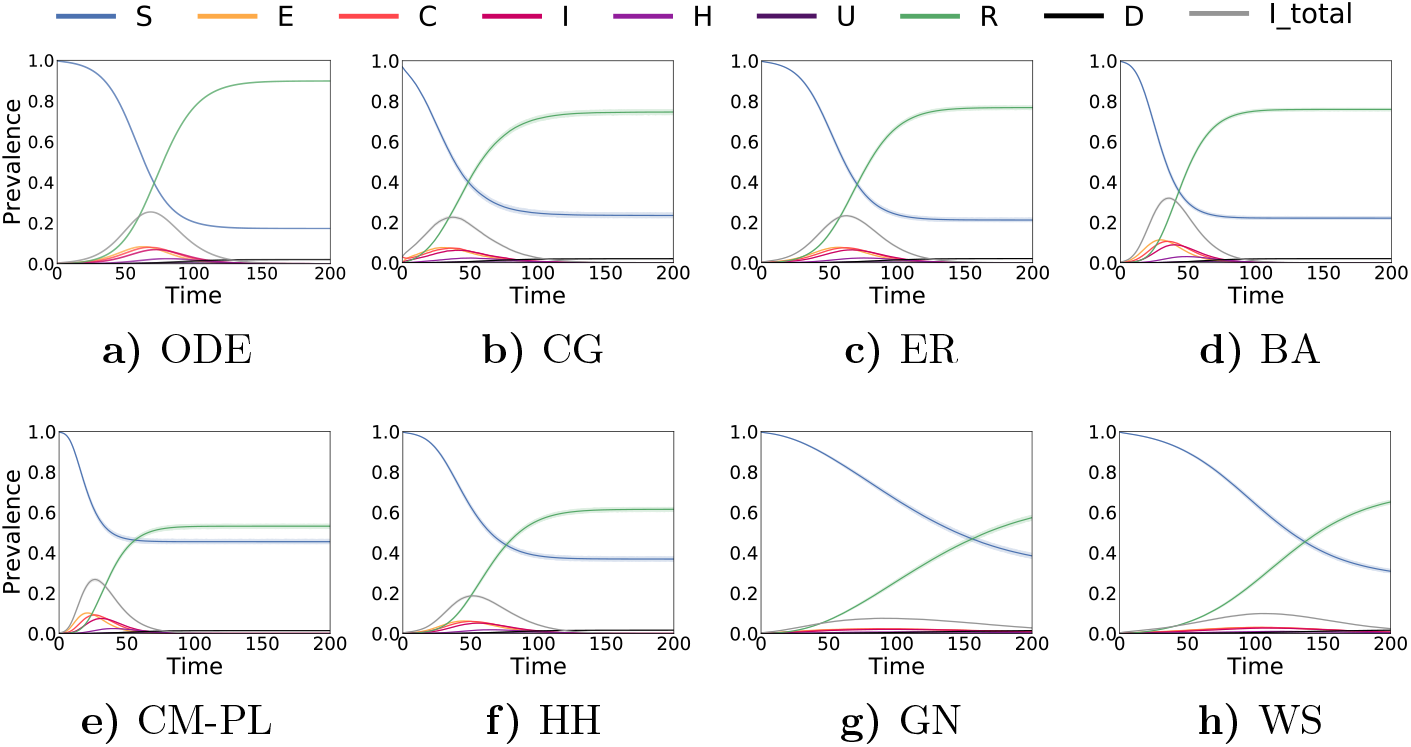
Exp. 1: Evolution of the mean fractions in each compartment over time with 95% confidence intervals (barely visible).

In contrast, in networks that admit super-spreaders (CM-PL, HH, and also BA), it is principally possible for *R_t_* to increase. For the CM-PL network, we have a very early and intense peak of the infection while the number of individuals ultimately affected by the virus (and consequently the death toll^5^) remains comparably small (when we remove the super-spreaders from the network while keeping the same *R*_0_, the death toll and the time point of the peak increase, plot not shown). Note that the high value of *R_t_* in Fig. 6c in the first days results from the fact that super-spreaders become exposed, which later infect a large number of individuals. As there are very few super-spreaders, they are unlikely to be part of the seeds. However, due to their high centrality, they are likely to be one of the first exposed nodes, leading to an “explosion” of the epidemic. In HH-networks this effect is way more subtle but follows the same principle.

**Experiment 2: Calibrating 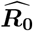 to a Fixed Peak**. Next, we calibrate λ such that each network admits an infection peak (regarding /total) of the same height (0.2). Results are shown in Fig. 7. They emphasize that there is no direct relationship between the number of individuals affected by the epidemic and the height of the infection peak, which is particularly relevant in the light of limited ICU capacities. It also shows that vastly different infection rates and basic reproduction numbers are acceptable when aiming at keeping the peak below a certain threshold.

**Fig. 5:**
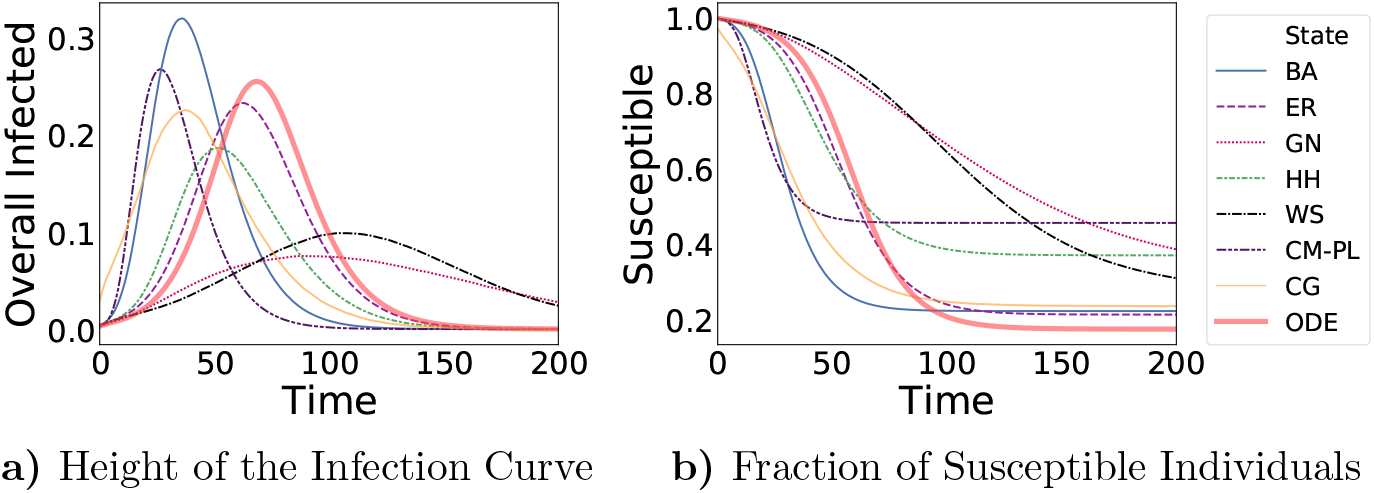
Exp. 1: Same data as in Fig. 4 but only the evolution Itotai and S are shown to highlight differences between networks.

**Fig. 6:**
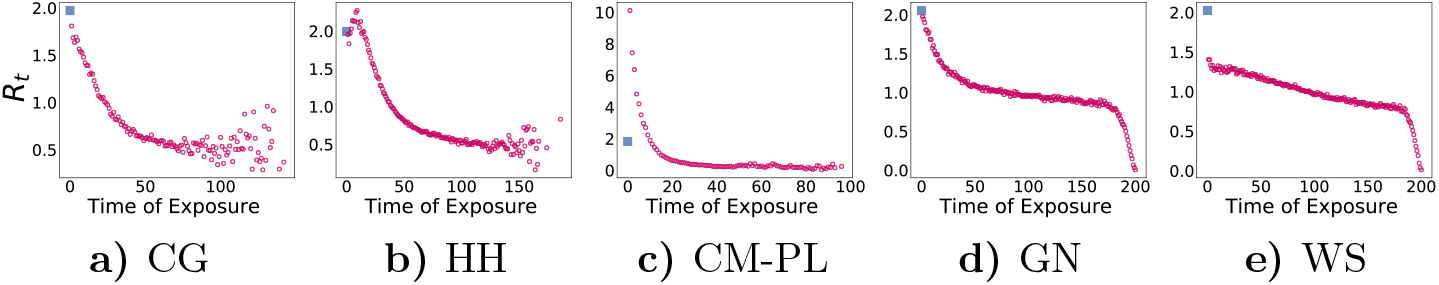
Exp. 1, Effective Reproduction Number: Evolution of the (mean) effective reproduction number, *R_t_*, over time, empirically evaluated. *x*-axis: Day at which a node becomes exposed, *y*-axis: (mean) number of neighbors this node infects while being a carrier or infected. Note that at later time points results are more noisy as the number of samples decreases. The first data-point is the simulation-based estimation of *R*_0_ and is shown as a blue square.

**Experiment 3: Sensitivity Regarding λ**. Assume we have an estimate of the infectiousness, λ, of COVID-19. How do changes of λ (e.g., by better hygiene) influence epidemic’s properties and what is the impact of uncertainty about the value? Here, we investigate how the height of the infection-peak and 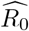 scale with λ for different topologies. Our results are illustrated in Fig. 8.

Noticeably, the relationship is concave for most network models but almost linear for the ODE model. This indicates that the networks models are more sensitive to small changes of λ (and *R*_0_). This suggests that the use of ODE models might lead to a misleading sense of confidence because, roughly speaking, it will tend to yield similar results when adding some noise to λ. That makes them seemingly robust to uncertainty in the parameters, while in reality the process is much less robust. Assuming that BA-networks resemble some important features of real social networks, the non-linear relationship between infection peak and infectiousness indicates that small changes of λ (which could be achieved through proper hand-washing, wearing masks, keeping distance, etc.) can significantly “flatten the curve”.

**Fig. 7:**
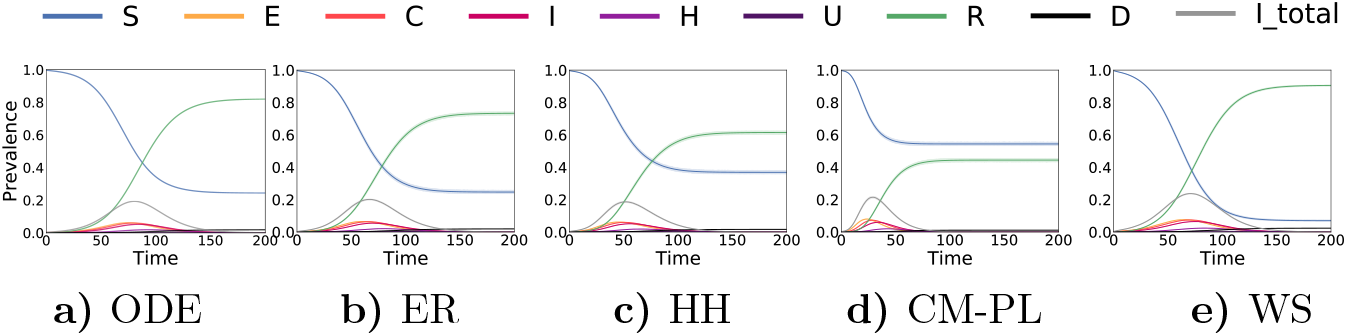
Exp. 2: Evolution of mean-fractions in each compartment over time with infectiousness calibrated such that the peak has the same height.

**Fig. 8:**
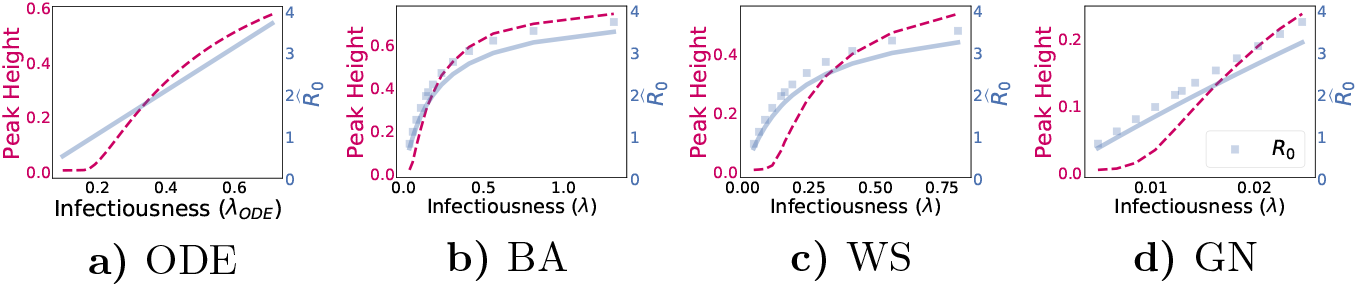
Exp. 3: 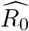 and maximal expected height of *I*_total_ (*expected* refers to samples, *maximal* refers to time) w.r.t. λ. For the network-models, *R*_0_ (given by Eq. (7)) is drawn as a scatter plot. Note the different scales on *x-*and *y*-axis

### 5.1 Discussion

In the series of experiments, we tested how various network types influence an epidemic’s dynamics. The network types highlight different potential features of real-world social networks. Most results do not contradict with real-world observations. For instance, we found that better hygiene and the truncation of super-spreaders will likely reduce the peak of an epidemic by a large amount. We also observed that, even when *R*_0_ is fixed, the evolution of *R_t_* largely depends on the network structure. For certain networks, in particular those admitting super-spreaders, it can even increase. An increasing reproduction number can be seen in many countries, for instance in Germany [21]. How much of this can be attributed to super-spreaders is still being researched. Note that super-spreaders do not necessarily have to correspond to certain individuals. It can also, on a more abstract level, refer to a type of events. We also observed that CM-PL networks have a very early and very intense infection peak. However, the number of people ultimately affected (and therefore also the death toll) remain comparably small. This is somewhat surprising and requires further research. We speculate that the fragmentation in the network makes it difficult for the virus to “reach every corner” of the graph while it “burns out” relatively quickly in region of the more central high-degree nodes.

## 6 Conclusions and Future Work

We presented results for a COVID-19 case study that is based on the translation of an ODE model to a stochastic network-based setting. We compared several interaction structures using contact graphs where one was (a finite version of) the implicit underlying structure of the ODE model, the complete graph. We found that inhomogeneity in the interaction structure significantly shapes the epidemic’s dynamic. This indicates that fitting deterministic ODE models to real-world data might lead to qualitatively and quantitatively wrong results. The interaction structure should be included into computational models and should undergo the same rigorous scientific discussion as other model parameters.

Contact graphs have the advantage of encoding various types of interaction structures (spatial, social, etc.) and they decouple the infectiousness from the connectivity. We found that the choice of the network structure has a significant and counterintuitive impact and it is very likely that this is also the case for the inhomogeneous interaction structure among humans. Specifically, networks containing super-spreaders consistently lead to the emergence of an earlier and higher peak of the infection. Moreover, the almost linear relationship between *R*_0_, λ_ODE_, and the peak intensity in ODE-models might also lead to misplaced confidence in the results. Regarding the network structure in general, we find that super-spreaders can lead to a very early “explosion” of the epidemic. Smallworldness, by itself, does not admit this property. Generally, it seems that—unsurprisingly—a geometric network is best at containing a pandemic. This is an indication for the effectiveness of corresponding mobility restrictions. Surprisingly, we found a trade-off between the height of the infection peak and the fraction of individuals affected by the epidemic in total.

For future work, it would be interesting to investigate the influence of non-Markovian dynamics. ODE-models naturally correspond to an exponentially distributed residence times in each compartment [49,17]. Moreover, it would be interesting to reconstruct more realistic contact networks. They would allow to investigate the effect of NPIs in the network-based paradigm and to have a well-founded scientific discussion about their efficiency. From a risk-assessment perspective, it would also be interesting to focus more explicitly on worst-case trajectories (taking the model’s inherent stochasticity into account). This is especially relevant because the costs to society do not scale linearly with the characteristic values of an epidemic. For instance, when ICU capacities are reached, a small additional number of severe cases might lead to dramatic consequences.

## Data Availability

Code is available on github

https://github.com/gerritgr/StochasticNetworkedCovid19

## Acknowledgements

We thank Luca Bortolussi and Thilo Krüger for helpful comments regarding the manuscript. This work was partially funded by the DFG project MULTIMODE.

## A Inferring *R*_0_ for Networks

**Fig. 9:**
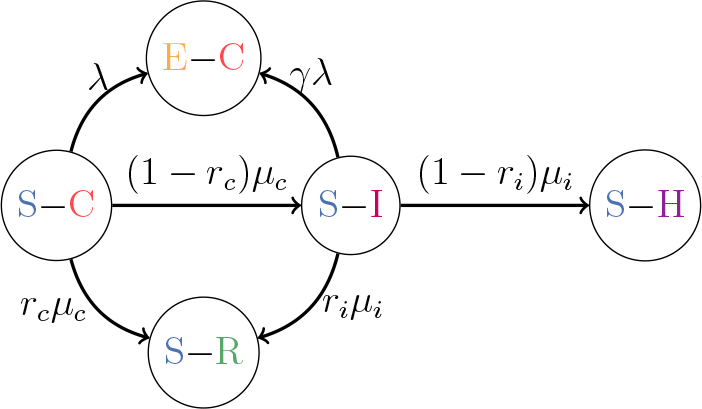
The reachability probability from S – C to E – C is related to *R*_0_. It expresses the probability that an infected node (patient zero, in C) infects a specific random (susceptible) neighbor. The infection can happen via two paths. Furthermore, we assume that this happens for all edges/neighbors of patient zero independently.

Assume a randomly chosen patient zero that is in compartment C. We are interested in *R*_0_ in the model given in Fig. 2 assuming *γ* > 0. Again, we consider each neighbor independently and multiply with *k*_mean_. Moreover, we have to consider the likelihood that patient zero infects a neighbor while being in compartment C and the possibility of transitioning to I and then transmitting the virus. This can be expressed as a reachability probability (cf. Fig. 9) and gives raise to the equation:

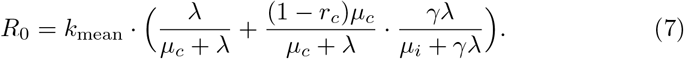

In the brackets, the first part of the sum expresses the probability that patient zero infects a random neighbor as long as it is in C. In the second part of the sum, the first factor expresses the probability that patient zero transitions to I before infecting a random neighbor. The second factor is then the probability of infecting a random neighbor as long as being in I. Note that, as we consider a fixed random neighbor, we need to condition the second part of the sum on the fact that the neighbor was not already infected in the first step.

## Footnotes

1 available at covidsim.eu

2 available at ddi.sutd.edu.sg

3 At the time of finalizing this manuscript, the model of Khailaie et al. seems to be updated in a similar way. However, it also became more complex (see gitlab.com/simm/covid19/secir/-/wikis/Report).

4 github.com/gerritgr/StochasticNetworkedCovid19

5 The number of fatalities in the figures is difficult to see, but it is (in the time limit) proportional to the number of recovered nodes.

## References

1. Arenas, A., Cota, W., Gomez-Gardenes, J., Gomez, S., Granell, C., Matamalas, J.T., Soriano-Panos, D., Steinegger, B.: Derivation of the effective reproduction number r for covid-19 in relation to mobility restrictions and confinement. medRxiv (2020)

2. Ball, F., Sirl, D., Trapman, P.: Analysis of a stochastic sir epidemic on a random network incorporating household structure. Mathematical Biosciences 224(2), 53–73 (2010)

3. Barrett, C.L., Beckman, R.J., Khan, M., Kumar, V.A., Marathe, M.V., Stretz, P.E., Dutta, T., Lewis, B.: Generation and analysis of large synthetic social contact networks. In: Proceedings of the 2009 Winter Simulation Conference (WSC). pp. 1003–1014. IEEE (2009)

4. Bi, Q., Wu, Y., Mei, S., Ye, C., Zou, X., Zhang, Z., Liu, X., Wei, L., Truelove, S.A., Zhang, T., et al.: Epidemiology and transmission of covid-19 in shenzhen china: Analysis of 391 cases and 1,286 of their close contacts. MedRxiv (2020)

5. Bistritz, I., Bambos, N., Kahana, D., Ben-Gal, I., Yamin, D.: Controlling contact network topology to prevent measles outbreaks. In: 2019 IEEE Global Communications Conference (GLOBECOM). pp. 1–6. IEEE (2019)

6. Bock, W., Adamik, B., Bawiec, M., Bezborodov, V., Bodych, M., Burgard, J.P., Goetz, T., Krueger, T., Migalska, A., Pabjan, B., et al.: Mitigation and herd immunity strategy for covid-19 is likely to fail. medRxiv (2020)

7. Buchholz, U., et al.: Modellierung von beispielszenarien der sars-cov-2-ausbreitung und schwere in deutschland (2020), (only available in German)

8. Chinazzi, M., Davis, J.T., Ajelli, M., Gioannini, C., Litvinova, M., Merler, S., y Piontti, A.P., Mu, K., Rossi, L., Sun, K., et al.: The effect of travel restrictions on the spread of the 2019 novel coronavirus (covid-19) outbreak. Science (2020)

9. Dehning, J., Zierenberg, J., Spitzner, F.P., Wibral, M., Neto, J.P., Wilczek, M., Priesemann, V.: Inferring covid-19 spreading rates and potential change points for case number forecasts. arXiv preprint arXiv:2004.01105 (2020)

10. Estrada, E., Knight, P.A.: A first course in network theory. Oxford University Press, USA (2015)

11. Ferguson, N., Laydon, D., Nedjati Gilani, G., Imai, N., Ainslie, K., Baguelin, M., Bhatia, S., Boonyasiri, A., Cucunuba Perez, Z., Cuomo-Dannenburg, G., et al.: Report 9: Impact of non-pharmaceutical interventions (npis) to reduce covid19 mortality and healthcare demand (2020)

12. Ferguson, N.M., Cummings, D.A., Fraser, C., Cajka, J.C., Cooley, P.C., Burke, D.S.: Strategies for mitigating an influenza pandemic. Nature 442(7101), 448–452 (2006)

13. Ghinai, I., Woods, S., Ritger, K.A., McPherson, T.D., Black, S.R., Sparrow, L., Fricchione, M.J., Kerins, J.L., Pacilli, M., Ruestow, P.S., et al.: Community transmission of sars-cov-2 at two family gatherings—chicago, illinois, february-march 2020 (2020)

14. Gleeson, J.P.: Binary-state dynamics on complex networks: Pair approximation and beyond. Physical Review X 3(2), 021004 (2013)

15. Grassly, N.C., Fraser, C.: Mathematical models of infectious disease transmission. Nature Reviews Microbiology 6(6), 477–487 (2008)

16. Grossmann, G., Backenkoehler, M., Wolf, V.: Importance of interaction structure and stochasticity for epidemic spreading: A covid-19 case study. ResearchGate (2020), https://www.researchgate.net/publication/341119247_Importance_of_Interaction_Structure_and_Stochasticity_for_Epidemic_Spreading_A_COVID-19_Case_Study

17. Großmann, G., Bortolussi, L., Wolf, V.: Rejection-based simulation of non-markovian agents on complex networks. In: International Conference on Complex Networks and Their Applications. pp. 349–361. Springer (2019)

18. Hackl, J., Dubernet, T.: Epidemic spreading in urban areas using agent-based transportation models. Future Internet 11(4), 92 (2019)

19. Hagberg, A., Swart, P., S Chult, D.: Exploring network structure, dynamics, and function using networkx. Tech. rep., Los Alamos National Lab.(LANL), Los Alamos, NM (United States) (2008)

20. Halloran, M.E., Ferguson, N.M., Eubank, S., Longini, I.M., Cummings, D.A., Lewis, B., Xu, S., Fraser, C., Vullikanti, A., Germann, T.C., et al.: Modeling targeted layered containment of an influenza pandemic in the united states. Proceedings of the National Academy of Sciences 105(12), 4639–4644 (2008)

21. Hamouda, O., et al.: Schätzung der aktuellen entwicklung der sars-cov-2-epidemie in deutschland-nowcasting (2020)

22. Hellewell, J., Abbott, S., Gimma, A., Bosse, N.I., Jarvis, C.I., Russell, T.W., Munday, J.D., Kucharski, A.J., Edmunds, W.J., Sun, F., et al.: Feasibility of controlling covid-19 outbreaks by isolation of cases and contacts. The Lancet Global Health (2020)

23. Holme, P.: Representations of human contact patterns and outbreak diversity in sir epidemics. IFAC-PapersOnLine 48(18), 127–131 (2015)

24. Huang, C., Liu, X., Sun, S., Li, S.C., Deng, M., He, G., Zhang, H., Wang, C., Zhou, Y., Zhao, Y., et al.: Insights into the transmission of respiratory infectious diseases through empirical human contact networks. Scientific reports 6, 31484 (2016)

25. Ioannidis, J.P.: Coronavirus disease 2019: the harms of exaggerated information and non-evidence-based measures. European journal of clinical investigation (2020)

26. Khailaie, S., Mitra, T., Bandyopadhyay, A., Schips, M., Mascheroni, P., Vanella, P., Lange, B., Binder, S., Meyer-Hermann, M.: Estimate of the development of the epidemic reproduction number rt from coronavirus sars-cov-2 case data and implications for political measures based on prognostics. medRxiv (2020)

27. Kiss, I.Z., Miller, J.C., Simon, P.L., et al.: Mathematics of epidemics on networks. Cham: Springer 598 (2017)

28. Kissler, S., Tedijanto, C., Goldstein, E., Grad, Y., Lipsitch, M.: Projecting the transmission dynamics of sars-cov-2 through the post-pandemic period (2020)

29. Klepac, P., Kucharski, A.J., Conlan, A.J., Kissler, S., Tang, M., Fry, H., Gog, J.R.: Contacts in context: large-scale setting-specific social mixing matrices from the bbc pandemic project. medRxiv (2020). https://doi.org/10.1101/2020.02.16.20023754, https://www.medrxiv.org/content/early/2020/03/05/2020.02.16.20023754

30. Lourenço, J., Paton, R., Ghafari, M., Kraemer, M., Thompson, C., Simmonds, P., Klenerman, P., Gupta, S.: Fundamental principles of epidemic spread highlight the immediate need for large-scale serological surveys to assess the stage of the sars-cov-2 epidemic. medRxiv (2020)

31. Machens, A., Gesualdo, F., Rizzo, C., Tozzi, A.E., Barrat, A., Cattuto, C.: An infectious disease model on empirical networks of human contact: bridging the gap between dynamic network data and contact matrices. BMC infectious diseases 13(1), 185 (2013)

32. Masuda, N., Holme, P.: Temporal network epidemiology. Springer (2017)

33. McCaw, J.M., Forbes, K., Nathan, P.M., Pattison, P.E., Robins, G.L., Nolan, T.M., McVernon, J.: Comparison of three methods for ascertainment of contact information relevant to respiratory pathogen transmission in encounter networks. BMC infectious diseases 10(1), 166 (2010)

34. Milne, G.J., Kelso, J.K., Kelly, H.A., Huband, S.T., McVernon, J.: A small community model for the transmission of infectious diseases: comparison of school closure as an intervention in individual-based models of an influenza pandemic. PloS one 3(12) (2008)

35. Nowzari, C., Preciado, V.M., Pappas, G.J.: Analysis and control of epidemics: A survey of spreading processes on complex networks. IEEE Control Systems Magazine 36(1), 26–46 (2016)

36. Pastor-Satorras, R., Castellano, C., Van Mieghem, P., Vespignani, A.: Epidemic processes in complex networks. Reviews of modern physics 87(3), 925 (2015)

37. Perez, L., Dragicevic, S.: An agent-based approach for modeling dynamics of contagious disease spread. International journal of health geographics 8(1), 50 (2009)

38. Prakash, B.A., Chakrabarti, D., Valler, N.C., Faloutsos, M., Faloutsos, C.: Threshold conditions for arbitrary cascade models on arbitrary networks. Knowledge and information systems 33(3), 549–575 (2012)

39. Preciado, V.M., Zargham, M., Enyioha, C., Jadbabaie, A., Pappas, G.: Optimal vaccine allocation to control epidemic outbreaks in arbitrary networks. In: 52nd IEEE conference on decision and control. pp. 7486–7491. IEEE (2013)

40. Prem, K., Liu, Y., Russell, T.W., Kucharski, A.J., Eggo, R.M., Davies, N., Flasche, S., Clifford, S., Pearson, C.A., Munday, J.D., et al.: The effect of control strategies to reduce social mixing on outcomes of the covid-19 epidemic in wuhan, china: a modelling study. The Lancet Public Health (2020)

41. Pung, R., Chiew, C.J., Young, B.E., Chin, S., Chen, M.I., Clapham, H.E., Cook, A.R., Maurer-Stroh, S., Toh, M.P., Poh, C., et al.: Investigation of three clusters of covid-19 in singapore: implications for surveillance and response measures. The Lancet (2020)

42. Rader, B., Scarpino, S., Nande, A., Hill, A., Dalziel, B., Reiner, R., Pigott, D., Gutierrez, B., Shrestha, M., Brownstein, J., Castro, M., Tian, H., Grenfell, B., Pybus, O., Metcalf, J., Kraemer, M.U.: Crowding and the epidemic intensity of covid-19 transmission. medRxiv (2020). https://doi.org/10.1101/2020.04.15.20064980, https://www.medrxiv.org/content/early/2020/04/20/2020.04.15.20064980

43. Riou, J., Althaus, C.L.: Pattern of early human-to-human transmission of wuhan 2019 novel coronavirus (2019-ncov), december 2019 to january 2020. Eurosurveillance 25(4) (2020)

44. Salathé, M., Kazandjieva, M., Lee, J.W., Levis, P., Feldman, M.W., Jones, J.H.: A high-resolution human contact network for infectious disease transmission. Proceedings of the National Academy of Sciences 107(51), 22020–22025 (2010)

45. Salehi, M., Sharma, R., Marzolla, M., Magnani, M., Siyari, P., Montesi, D.: Spreading processes in multilayer networks. IEEE Transactions on Network Science and Engineering 2(2), 65–83 (2015)

46. Sapiezynski, P., Stopczynski, A., Lassen, D.D., Lehmann, S.: Interaction data from the copenhagen networks study. Scientific Data 6(1), 1–10 (2019)

47. Soriano-Panos, D., Ghoshal, G., Arenas, A., Gómez-Gardenes, J.: Impact of temporal scales and recurrent mobility patterns on the unfolding of epidemics. Journal of Statistical Mechanics: Theory and Experiment 2020(2), 024006 (2020)

48. Stewart, W.J.: Probability, Markov chains, queues, and simulation: the mathematical basis of performance modeling. Princeton university press (2009)

49. Van Mieghem, P., Van de Bovenkamp, R.: Non-markovian infection spread dramatically alters the susceptible-infected-susceptible epidemic threshold in networks. Physical review letters 110(10), 108701 (2013)

50. Vynnycky, E., White, R.: An introduction to infectious disease modelling. OUP oxford (2010)

51. Wilson, N., Barnard, L.T., Kvalsig, A., Verrall, A., Baker, M.G., Schwehm, M.: Modelling the potential health impact of the covid-19 pandemic on a hypothetical european country. medRxiv (2020)

52. Wilson, N., Barnard, L.T., Kvalsvig, A., Baker, M.: Potential health impacts from the covid-19 pandemic for new zealand if eradication fails: Report to the nz ministry of health (2020)

